# Google Trends as a predictive tool for COVID-19 vaccinations in Italy: a retrospective infodemiological analysis

**DOI:** 10.1101/2021.11.29.21267012

**Authors:** Alessandro Rovetta

## Abstract

**Background:** Google Trends is an infoveillance tool widely used by the scientific community to investigate different user behaviors related to COVID-19. However, several limitations regarding its adoption are reported in the literature.

**Objective:** This brief paper aims to provide an effective and efficient approach to investigating vaccine adherence against COVID-19 via Google Trends.

**Methods:** Through the cross-correlational analysis of well-targeted hypotheses, we investigate the predictive capacity of web searches related to COVID-19 towards vaccinations in Italy from November 2020 to November 2021. The keyword “vaccine reservation” (VRQ) was chosen as it reflects a real intention of being vaccinated (V). Furthermore, the impact of the second-largest Italian national newspaper on vaccines-related web searches was investigated to evaluate the role of the mass media as a confounding factor.

**Results:** Simple and generic keywords are more likely to identify the actual web interest in COVID-19 vaccines than specific and elaborated keywords. Cross-correlations between VRQ and V were very strong and significant (min r^2^ = .460, P<.001, lag = 0 weeks; max r^2^ = .903, P < .001, lag = 6 weeks). Cross-correlations between VRQ and news about COVID-19 vaccines have been markedly lower and characterized by greater lags (min r^2^ = .190, P=.001, lag = 0 weeks; max r^2^ = .493, P < .001, lag = -10 weeks). No correlation between news and vaccinations was sought since the lag would have been too high.

**Conclusions:** This research provides strong evidence in favor of using Google Trends as a surveillance and prediction tool for vaccine adherence against COVID-19 in Italy. These findings prove that the search for suitable keywords is a fundamental step to reduce confounding factors. Additionally, targeting hypotheses helps diminish the likelihood of spurious correlations. It is recommended that Google Trends be leveraged as a complementary infoveillance tool by government agencies to monitor and predict vaccine adherence in this and future crises by following the methods proposed in this manuscript.

## Introduction

Google Trends has often been employed by the scientific community to conduct infodemiological and epidemiological analyzes [1, 2]. However, some authors have shown severe limitations in its use as a surveillance tool, including anomalies in results and mass media influence [3, 4]. Nonetheless, various strategies have been proposed in the literature to address these weaknesses [4, 5]. Taking the latter into account, in this brief paper, Google Trends is used to investigate vaccine adherence in Italy against COVID-19. Indeed, COVID-19 vaccines are essential to contain the infection, limiting the spread of new variants of concern and drastically reducing the severity of the disease [6]. Furthermore, the use of effective and efficient infoveillance techniques is also necessary for any future health crises. Therefore, this research proposes an approach capable of targeting the hypotheses and eliminating the anomalies of Google Trends, thus reducing the likelihood of running into spurious correlations and having statistically uncertain outcomes. Specifically, the ability to predict the COVID-19 vaccination trend in Italy based on web queries relating to the booking of the COVID-19 vaccination is examined.

## Methods

### Procedure summary

The hypothesis to be verified is that the “COVID-19 vaccine reservation” queries, abbreviated in VRQ, can predict the trends of national and regional vaccinations (V). To achieve this scope, cross-correlations between VRQ and V were searched. Only “interest over time” datasets were analyzed to avoid the problem of RSV anomalies. To quantify the impact of mass media on RSV, cross-correlations between VQR and the COVID-19 vaccines-related headlines of the second most read newspaper in Italy, “La Repubblica,” were searched. In particular, “La Repubblica” was chosen both for its large readership and its online historical database (which allows the user to easily search for published articles containing a list of specific keywords).

### Data collection

The keyword “prenotazione vaccino” (vaccine reservation), was selected since it clearly expresses the desire to administer the dose of a vaccine. The goodness of VRQ in identifying the web interest in COVID-19 vaccine queries is reported in the Results section. The Google Trends parameters have been set as follows: region = Italy, period = 1 November 2020 – 27 November 2021, category = All categories, search type = Web Search. To perform a historical analysis of the timeseries, the “period” parameter has been changed to “Past 5 years.” To analyze regional trends, the “region” parameter was changed from “Italy” to “[the name of the region concerned].” The “interest over time” datasets were downloaded in “.csv format.” Regarding national vaccinations, the dataset was downloaded from the “Github” platform [7]. The keyword “vaccino, vaccini, astrazeneca, pfizer, moderna, johnson&johnson, vaxzevria, comirnaty, pikevax” was searched for in the historical archive of the newspaper “La Repubblica” [8]. In particular, the number of articles containing the aforementioned keyword were counted from week to week until covering the period November 2020 - November 2021. The filter has been set to “ricerca avanzata” (advanced search) and “almeno una [parola]” (at least one [word]).

### Statistical analysis

We verified the shape of the data distribution both graphically and through the Shapiro-Wilk test. Since the datasets were not normal (P<.001) and we were not interested in looking for above or below threshold correlations, we adopted the Spearman correlation [9]. To check the discrepancy between two timeseries, we exploited quantifiers such as percentage difference (used to compare two simultaneous series and indicated with “δ”) and percentage increase (used to compare two consecutive series and indicated with “Δ”). The statistical significance of the discrepancies between average values was measured through the Welch t-test (t), which is also valid for large non-normal datasets [10, 11]. When two contiguous timeseries were compared, a graphic check was carried out to guarantee the absence of seasonalities and trends capable of compromising the result. No correlation between vaccine-related news and vaccinations was sought since the lag would have been too high (see Figure 2 for details).

## Results

The adoption of the “vaccine reservation” query (VRQ) for our purpose is validated by the very strong correlation with the “covid vaccine” and “vaccine” queries (Figure 1) and the marked increase of its relative search volume in the period November 2020 - November 2021 compared to the past four years (Δ=11,500%, t=6.8, P<.001).

**Figure 1.**
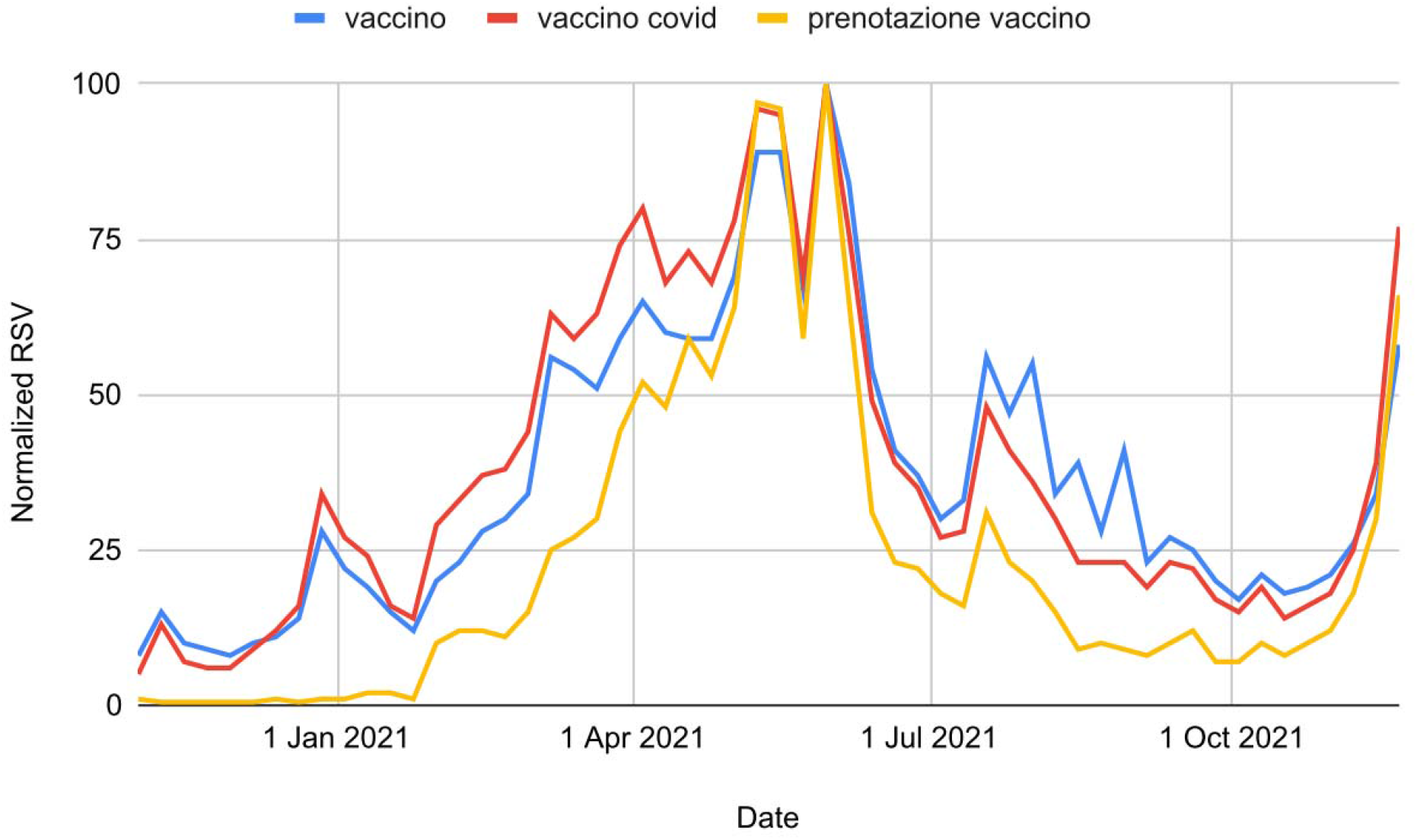
Normalized relative search volumes of the queries: “vaccino” (vaccine), “vaccino covid” (covid vaccine), and “prenotazione vaccino” (vaccine reservation).

**Figure 2.**
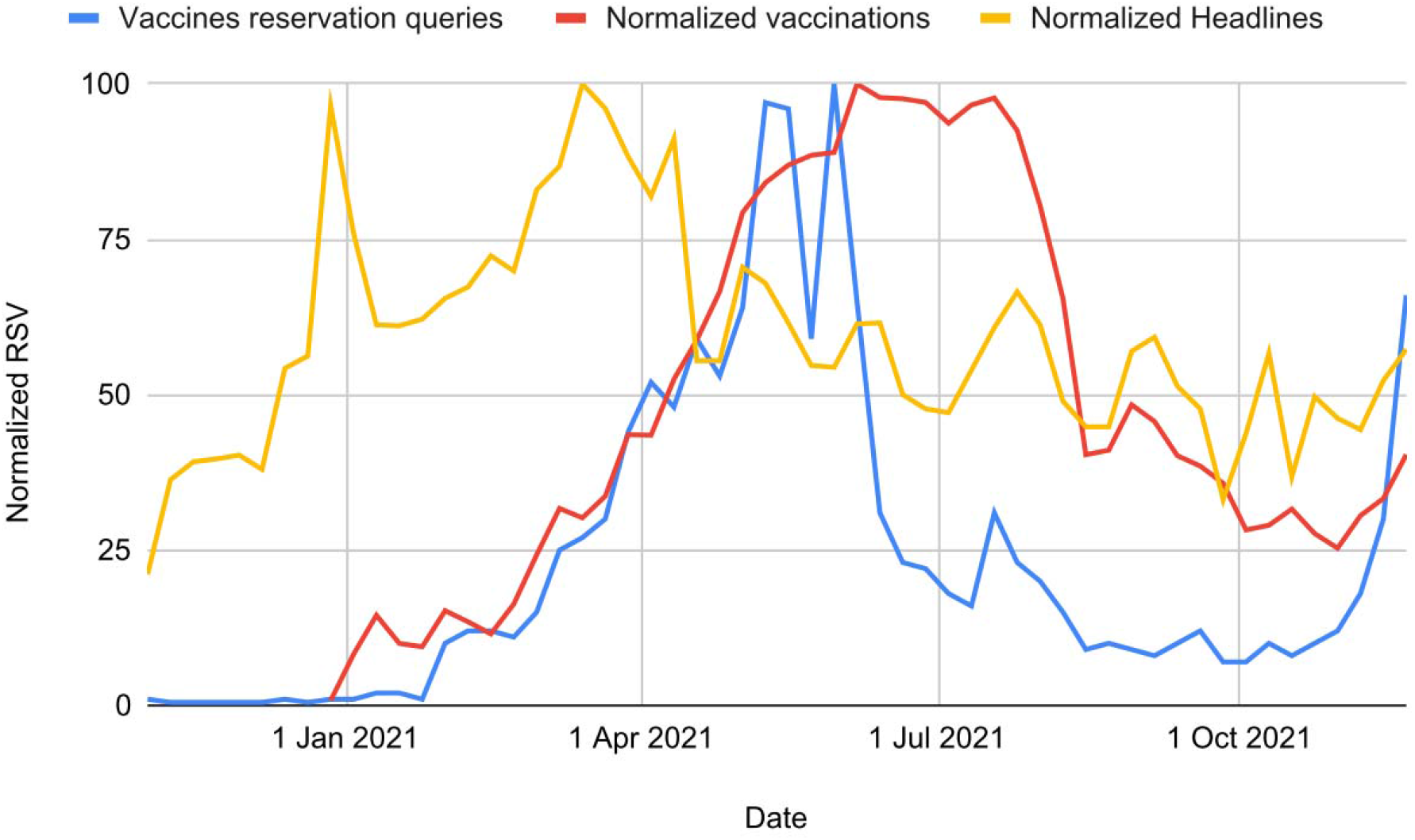
Comparison between the vaccines-related articles of the newspaper “La Repubblica,” national vaccinations, and national queries on vaccination reservations from November 2020 to November 2021.

VRQ’s relative search volume has significantly exceeded that of searches for specific names such as “pfizer reservation,” “astrazeneca reservation,” “moderna reservation,” and “johnson&johnson reservation” (δ = 190%, t = 6.6, P < .001). Table 1 shows very strong correlations between VRQ and the national vaccination (V) trends (min r^2^ = .460, P<.001, lag = 0 weeks; max r^2^ = .903, P < .001, lag = 6 weeks). Significant correlations were also highlighted between VRQ’s and the vaccines-related headlines of the newspaper “La Repubblica” (Table 2). However, in this case, the lags were greater, and the correlations were markedly lower (min r^2^ = .190, P=.001, lag = 0 weeks; max r^2^ = .493, P < .001, lag = -10 weeks). The comparison of the trends is shown in Figure 2. Figure 3 shows the trend of the VRQ in all Italian regions. In particular, it can be observed that all trends have been similar. Furthermore, Figure 3 is compatible with vaccination trends at the regional level [12].

**Table 1.**
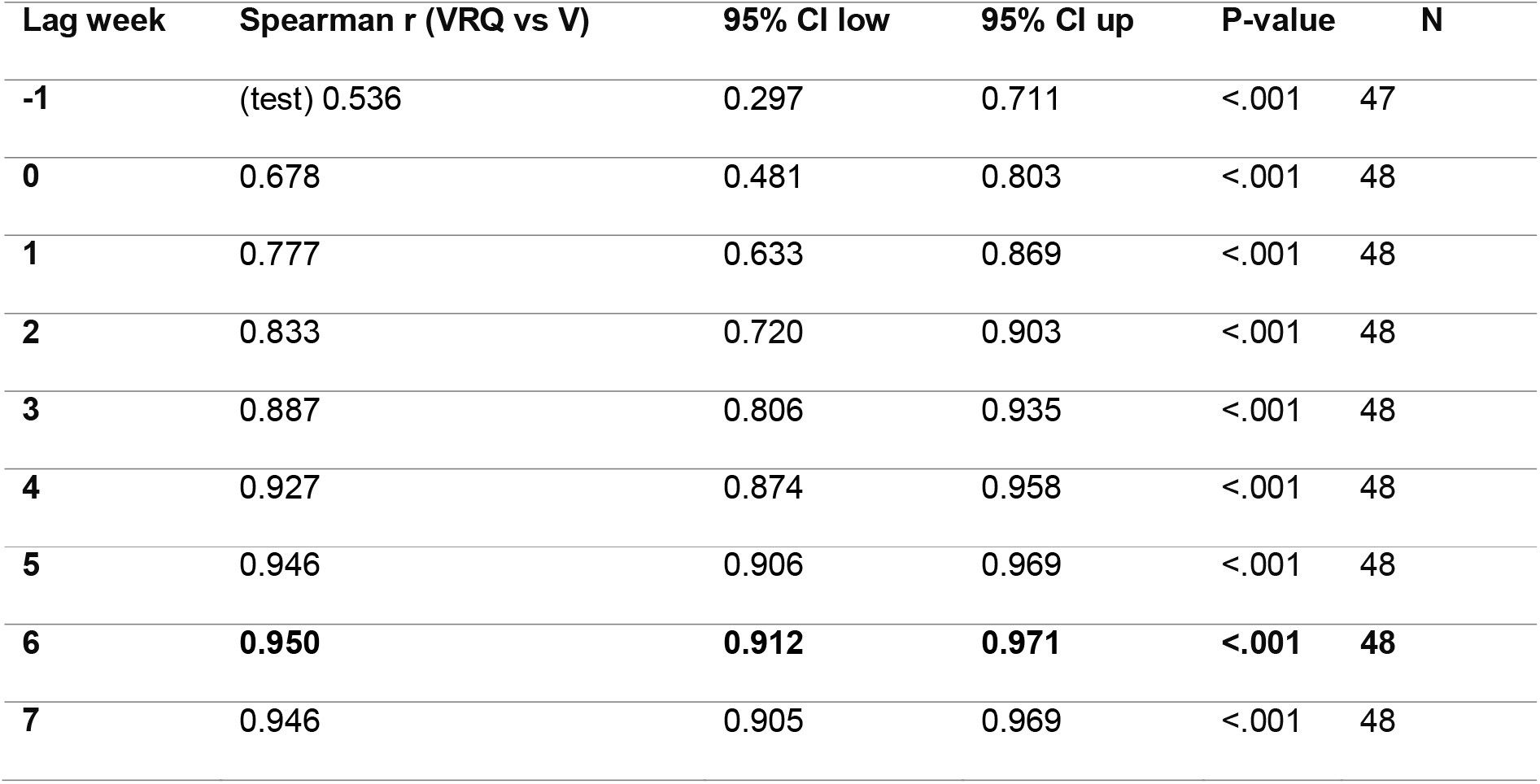
Cross-correlations between the “vaccine reservation” query and vaccination administrations in Italy between November 2020 and November 2021.

**Table 2.** Cross-correlations between the “vaccine reservation” query (VRQ) and “La Repubblica” vaccines-related headlines (VRH) between November 2020 and November 2021.

| Lag week | Spearman r (VRH vs VRQ) | 95% CI low | 95% CI up | P-value | N |
| --- | --- | --- | --- | --- | --- |
| -11 | 0.686 | 0.492 | 0.815 | <.000 | 45 |
| -10 | **0.702** | **0.518** | **0.824** | **<.001** | **46** |
| -9 | 0.700 | 0.518 | 0.822 | <.000 | 47 |
| -8 | 0.673 | 0.482 | 0.803 | <.001 | 48 |
| -7 | 0.660 | 0.466 | 0.793 | <.001 | 49 |
| -6 | 0.657 | 0.464 | 0.790 | <.001 | 50 |
| -5 | 0.580 | 0.363 | 0.737 | <.001 | 51 |
| -4 | 0.517 | 0.285 | 0.692 | <.001 | 52 |
| -3 | 0.487 | 0.250 | 0.668 | <.001 | 53 |
| -2 | 0.464 | 0.224 | 0.650 | .001 | 54 |
| -1 | 0.452 | 0.213 | 0.640 | .001 | 55 |
| 0 | 0.436 | 0.196 | 0.627 | .001 | 56 |

**Figure 3.**
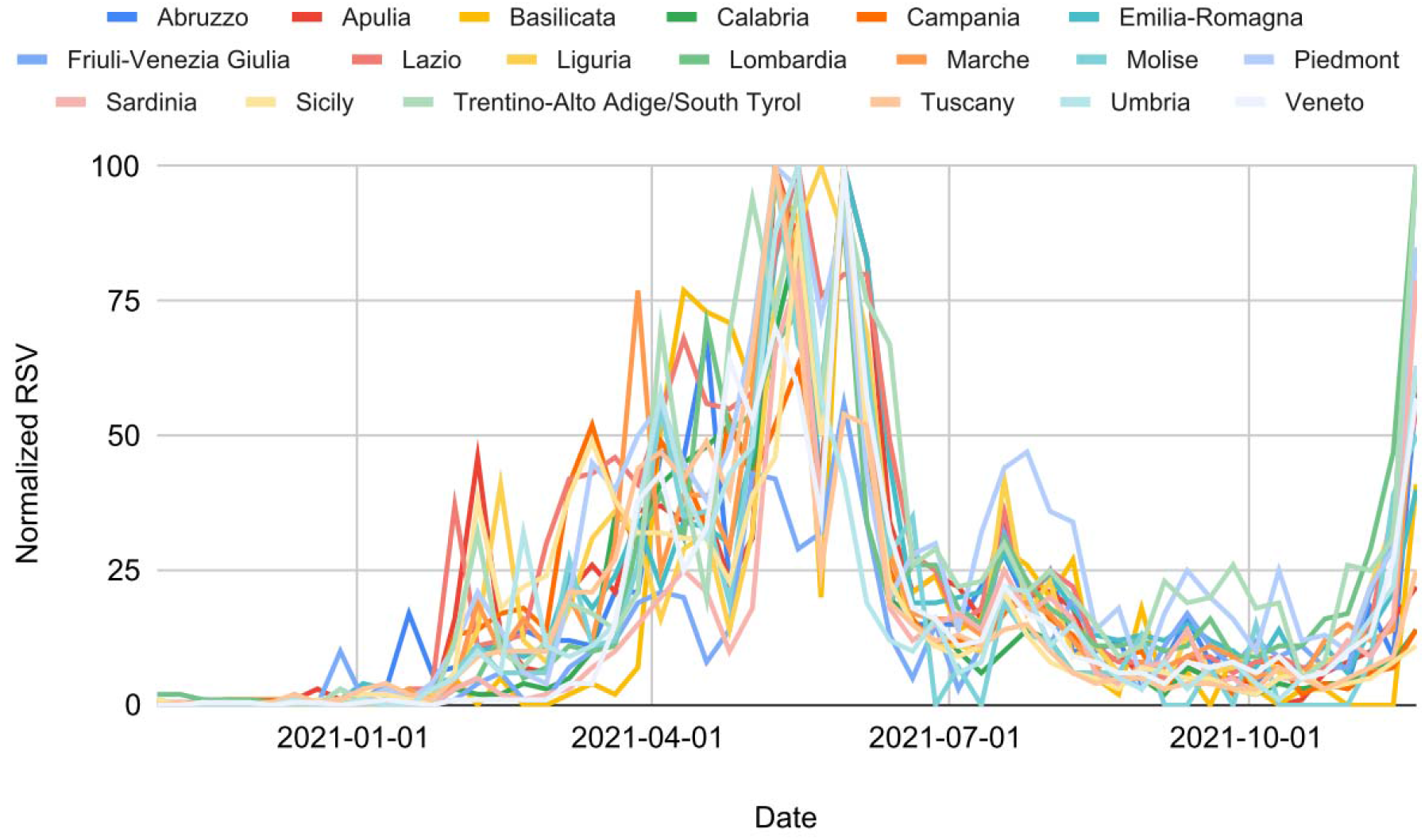
Comparison between the “prenotazione vaccino” (vaccine reservation) queries of all Italian regions from November 2020 to November 2021.

## Discussion

This study shows a marked and significant cross-correlation between the web desire to be vaccinated and vaccinations against COVID-19 in Italy. Based on the lower cross-correlations between vaccine-related news and vaccine web searches, the mass media may have only partially influenced web searches related to vaccine booking. Nevertheless, even assuming an impact of the mass media on these queries, this does not compromise the adoption of Google Trends as a predictive tool for vaccinations: indeed, the mass media could push users to search for online information on vaccines and then book their administration. Furthermore, COVID-19 vaccine reservation is easily obtainable through a user-friendly online procedure proposed by the regional health organizations (e.g., [13]). Therefore, it is likely that the cross-correlations found between vaccine-related queries and vaccinations are not spurious. Alongside this, it is necessary to consider that the Italian mass media have even risked compromising the effectiveness of the vaccination campaign against COVID-19 by providing infodemic news on very rare side effects [14]. Hence, it is plausible that, given the high number of vaccinations achieved at the national level, more authoritative sources have also been consulted by users. The capacity to provide accurate predictions on vaccination trends several weeks in advance is an extremely relevant epidemiological tool for developing future containment strategies [15]. These findings show that Google Trends can be exploited for this purpose if used properly. The search for simple well-targeted keywords on Google Trends is more likely to return the actual scenario of web interest on COVID-19 vaccines. Specifically, it is essential not to use too complex or specific names - which tend to be ignored by users - and try to express a precise action (in this case, the vaccine reservation).

Among the limitations of this paper, it is fair to emphasize that no definitive causal evidence has been provided and unknown confounders may have skewed the results in unpredictable ways. Moreover, the variability of time-lags between online booking and vaccine administration was not considered in this study.

## Data Availability

All data produced in the present work are contained in the manuscript.

